# Correlates of protection, thresholds of protection, and immunobridging in SARS-CoV-2 infection

**DOI:** 10.1101/2022.06.05.22275943

**Authors:** David S Khoury, Timothy E Schlub, Deborah Cromer, Megan Steain, Youyi Fong, Peter B Gilbert, Kanta Subbarao, James A Triccas, Stephen J Kent, Miles P Davenport

## Abstract

Several studies show neutralizing antibody levels are an important correlate of immune protection from COVID-19 and have estimated the relationship between neutralizing antibodies and protection. However, a number of these studies appear to yield quite different estimates of the level of neutralizing antibodies required for protection. Here we show that after normalization of antibody titers current studies converge on a consistent relationship between antibody levels and protection from COVID-19.

## Introduction

Determining the relationship between immune response and protection from symptomatic SARS-CoV-2 infection (i.e. COVID-19) is important to predicting the future effectiveness of vaccines. It should allow ‘immunobridging’ (i.e. predicting the efficacy of candidate vaccines) to facilitate the approval of new or updated vaccines based on immunogenicity data, without the need for large phase 3 trials (*1*). Immunobridging is currently used for approval of seasonal influenza vaccines in the European Union and US and both reduces the costs and the time required for vaccine development. In addition, defining levels of immunity required for protection from novel SARS-CoV-2 variants will be an important consideration in predicting population level immunity to infection and guiding public health policy on vaccination and boosting.

Several studies have shown that higher neutralizing antibody levels are associated with immune protection from symptomatic SARS-CoV-2 infection during short-term follow-up post vaccination (*2-6*). In addition, three of these studies also tried to estimate the level of protection associated with particular antibody levels (*2-4*). These studies used two different approaches to estimate the relationship between neutralizing antibody levels and vaccine efficacy (Figure 1)(for simplicity we refer to this relationship as the ‘protection curve’, see Glossary). Although these studies have reported ‘threshold’ antibody levels required for 50% or 70% protection, all studies found that protection changes gradually with neutralization titer, and thus there was not a strict threshold below which individuals are not protected or above which protection is achieved.

**Figure 1:**
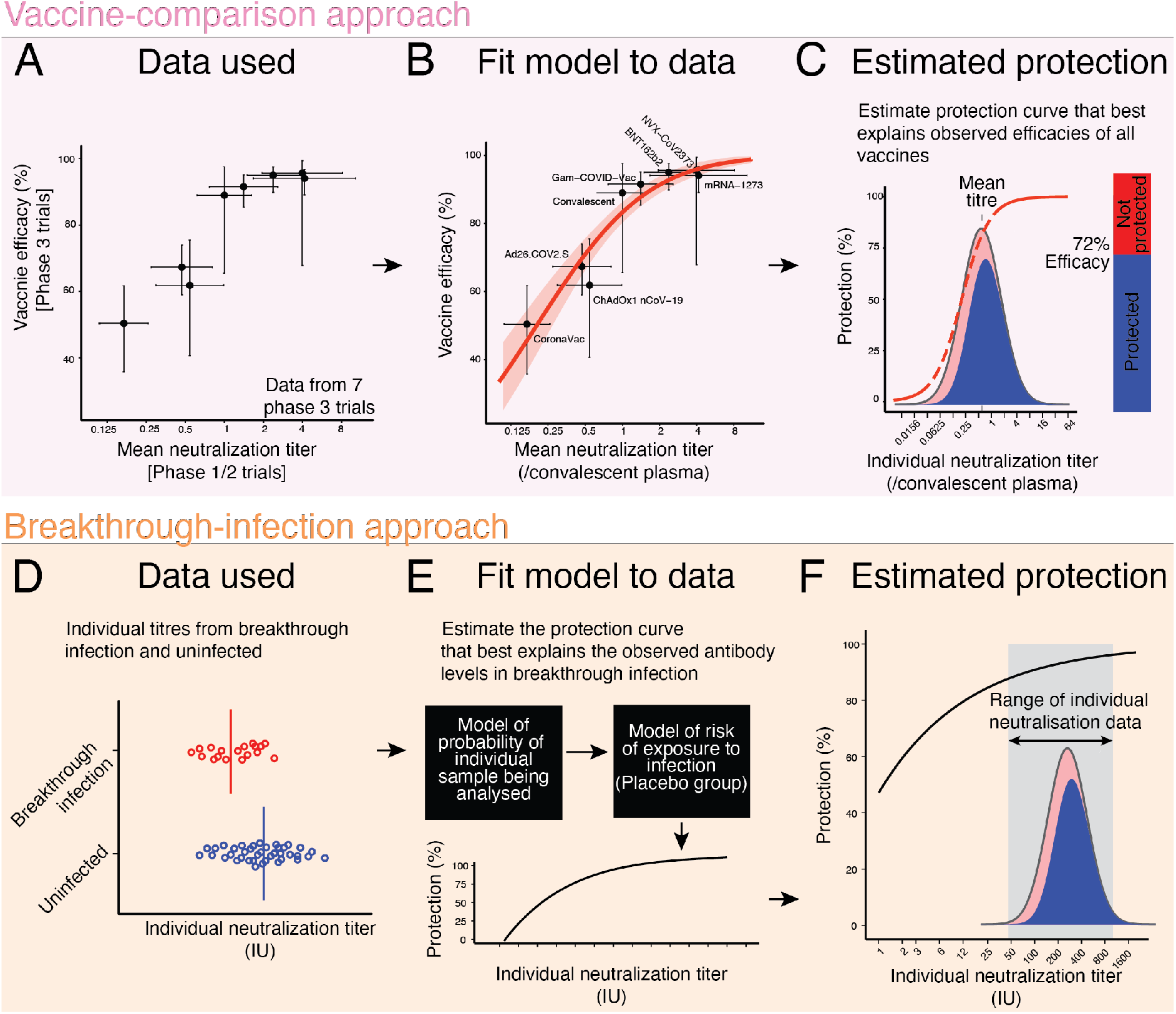
Predicting protection from symptomatic SARS-CoV-2 infection: Two different approaches have been employed thus far to understand the relationship between neutralizing antibody titers and protection from COVID-19 (we term this relationship the ‘protection curve’). Here, the vaccine-comparison (A-C) and breakthrough-infection (D-F) approaches to estimating the ‘protection curve’ are illustrated schematically. In the vaccine-comparison approach, data on mean neutralization titer from phase 1 /2 vaccine trials (normalized to convalescent subjects in the same study)(x-axis) and observed vaccine efficacy against symptomatic SARS-CoV-2 infection in phase 3 trials is used (y-axis, n=7 vaccine trials included, and one study of infection risk in convalescent individuals). Using the observed distribution in neutralization titers for a given vaccine, the protection curve is used to predict the proportion susceptible (blue) or protected (red) at different neutralizing antibody levels. By fitting across all vaccines and convalescent subjects simultaneously (panel B), one can derive the protection curve that best fits the neutralization and protection data (panel C). (D) The breakthrough-infection model uses neutralization titers of subjects with symptomatic breakthrough infections (n = 36 and n = 47 for mRNA-1273 and ChAdOx1 nCoV19 respectively) and uninfected individuals (n = 1005 and n = 828, respectively)(*3, 4*). The breakthrough-infection modeling uses an underlying ‘risk model’ to adjust for demographic risk factors and adjusted for the probability of being sampled in the study to remove these potential sources of bias (E). The protection curve then reflects an estimate of the vaccine efficacy in subgroups of individuals with specific neutralization titers after adjusting for the two-phase sampling design (F).

The first study of immune correlates by Khoury et al. used a ‘vaccine-comparison’ approach by fitting the relationship between mean neutralizing antibody levels (in phase 1 / 2 trials) and observed vaccine efficacy (in phase 3 trials) across seven vaccines and convalescent individuals (normalizing neutralization titers to convalescent subjects in each study)(*2*) (Figure 1A-C). This study estimated that the neutralizing antibody level associated with 50% protection from COVID-19 was around 20% of the mean convalescent titer (or 54 international units (IU)/ml)(*2*). More recently, two studies compared neutralizing antibody titers from mRNA 1273 or ChAdOx1 nCoV19-vaccinated subjects with or without symptomatic ‘breakthrough infection’ (Figure 1D-F). These studies reported 70% protective thresholds ranging from 4 IU/ml to 33 IU/ml depending on the assay used, suggesting a potential role of assay differences in these discrepancies (see full discussion in supplementary material) (*3, 4*).

### Reconciling the studies on thresholds of protection

A major limitation for the field is the lack of a standardized assay to measure in vitro neutralization titers. Although an International Standard has been established (*7*), the reported titers seem very affected by the assay used. For example, even against the same stocks of pooled convalescent plasma (for example the WHO 20/130 standard), different assays produced geometric mean neutralization titers (GMT) that varied from 120 to >12000 (table 7 in reference (*7*)). Even after standardization of different laboratories’ assays into international units, there was still up to a 50-fold difference in the reported neutralization titer of another standardized sample across the different assays (Table 8 from reference (*7*)). This difference in neutralization titers across different assays is also evident in comparing the three studies quantifying the threshold of protection (*2-4*). For example, in Gilbert et al. the GMT for mRNA-1273 is reported as ∼247 IU/ml (*4*), compared to a GMT for mRNA-1273 of 1057 IU/ml in Khoury et al (See supplementary material for details of assays and conversion to IU). A quick survey of the literature reveals six estimates of the GMT for the mRNA-1273 vaccine reported in IU/mL. These ranged between 247 IU/mL (95% CI: 231-264) and 1404 (95% CI: 795-2484) IU/mL depending on the study (Table S1). Similarly, estimates of the GMT of ChAdOx1 nCoV19 vaccinees range from 23 IU/ml (*3*) to 144 IU/ml (estimated in (*2*) from (*8*)). It is clear from these discrepancies that expression of titers in international units is insufficient to normalize between assays and to compare the thresholds of protection reported in these studies, and that this is likely due to differences in the assays themselves (*9*).

An alternative approach to normalizing neutralization titers between studies is to assume that similar groups (of vaccinees) should have similar titers. For example, rather than relying on conversion to the WHO international units, we can assume that the mean neutralization for the mRNA-1273 vaccinees is similar in the phase 1 /2 trial (as analysed in Khoury et al (*2, 10*)) and in the phase 3 trial (as analysed in Gilbert et al (*4, 11*)). This normalization is limited because it does not account for differences in baseline characteristics of the cohort vaccinated in each study, such as age, which may influence neutralization titers. However, since immunobridging studies also rely on comparison of vaccine titers in different groups, this is a pragmatic approach to overcoming the limitations of comparing between different assays.

Applying this normalization approach allows us to compare the ‘protection curves’ across different studies. In Figure 2 we align the data by assuming the mean titer for mRNA-1273-vaccinated or ChAdOx1 nCoV19-vaccinated subjects is the same between the phase 1 / 2 studies and the phase 3 studies for each vaccine (detailed methods in supplementary material). Although this normalization is independent of the x-axis scale used, we plot both the curves onto a ‘fold-of-convalescent’ level scale as developed in Khoury et al (*2*) for illustration. This transformation now allows a more direct comparison of the protection curve across the three studies. Considering the mRNA-1273 breakthrough infection model (*4*)(Figure 2A) for example, we see that it has good agreement with the Khoury et al. model at the higher neutralization levels achieved with mRNA-1273 vaccination (albeit a seemingly slightly lower maximum protection level predicted in the breakthrough infection model), but very poor agreement at low neutralization levels. This is easily understandable when one considers the distribution of individual neutralization titers in the mRNA-1273 breakthrough infection study, with only approximately 10% of participants having a neutralization titer less than the average of early convalescent subjects (Figure 2A). Thus, there are sparse neutralization data with which to estimate protection at lower neutralization levels (hence the wide confidence bands in this region of the curve). Similarly, the ChAdOx1 nCoV19 protection curve (Figure 2B) shows good agreement with the Khoury et al. analysis in the region where neutralization data is available in the breakthrough-infection study (Figure 2B).

**Figure 2:**
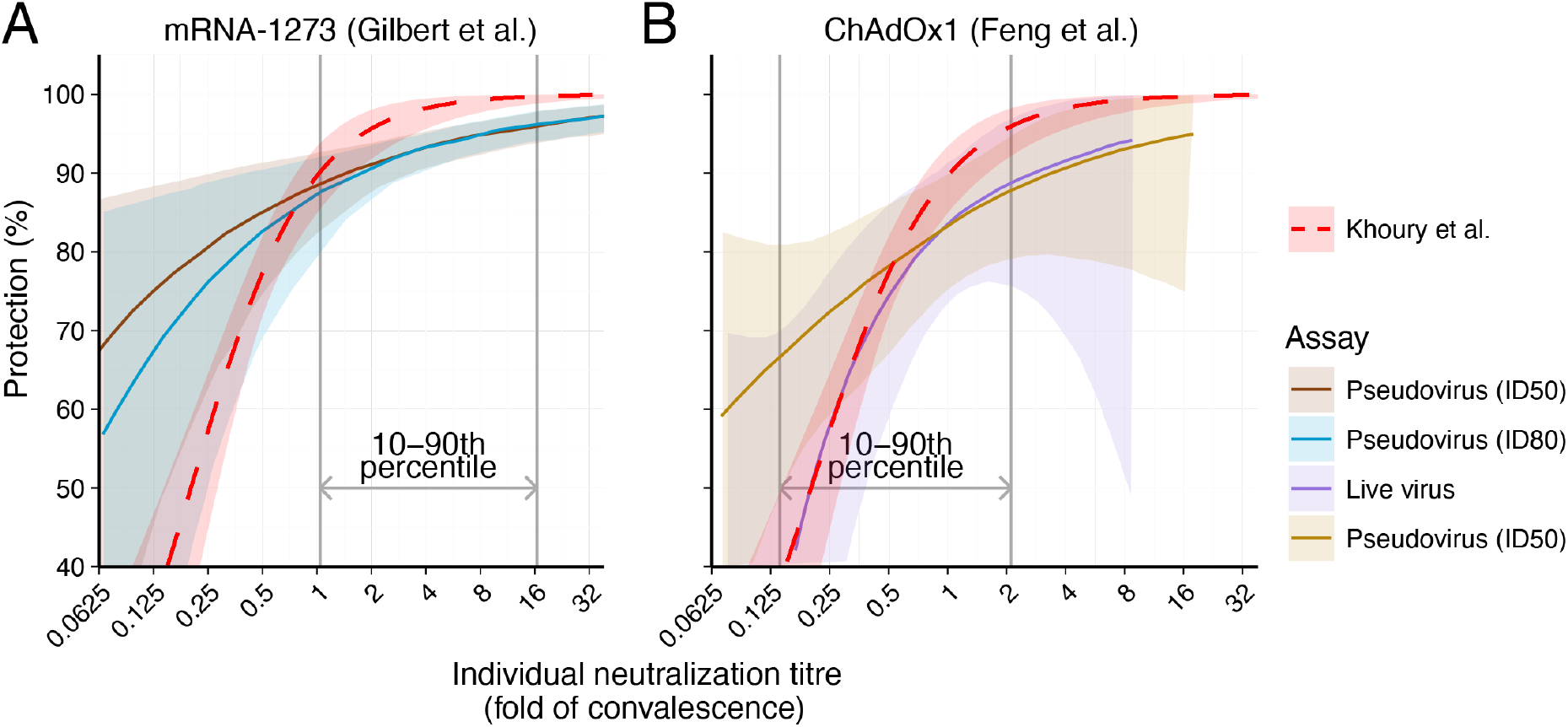
Comparing the estimated protection curves: The relationships between vaccine efficacy against COVID-19 infection (y-axis) and neutralization titers that were estimated in each study are shown (protection curve). The protection curve derived from the vaccine-comparison model is shown (red dashed line), and is compared with the modeled protection curves estimated from breakthrough infection studies by Gilbert et al. (dark brown and teal lines, for the results from the 50% and 80% neutralization titer in in vitro pseudovirus assays, ID50 and ID80, respectively) (A) and Feng et al. (purple and light brown, for the results from in vitro native (live) SARS-CoV-2 virus and pseudovirus neutralization assays, respectively) (B). Shaded areas are the 95% CI from each model. These curves were extracted from the studies (see supplementary material), and differences in assays were controlled for by normalizing the curve from each study by the mean neutralization titer of the uninfected vaccinees in each study. These normalized curves were then represented on a fold-of-convalescent scale by multiplying by the mean neutralization titer of vaccinees compared with convalescent subjects as reported in the Phase 1/2 trials(*8, 10*). We note that the vaccine-comparison model agrees closely with the breakthrough-infection models in the neutralization titer ranges where data were most abundant (vertical gray lines indicate 10-90th percentiles of the data available in each study respectively).

The broad confidence intervals and divergence of the models where neutralization data are sparse suggests caution in extrapolating the relationship between neutralization and protection beyond the ranges of data available in each study. The vaccine comparison approach has the advantage of fitting to a large span of neutralization titers (a 20-fold range in GMT between the seven vaccines (*2*)), allowing prediction of the vaccine efficacy over a wide range of neutralization titers. Importantly, since none of the reported phase 3 studies of ancestral SARS-CoV-2 infection has reported an efficacy below 50% or above 95%, the vaccine-comparison analysis also extrapolates efficacy above and below these levels. However, studies of vaccine efficacy and effectiveness against the SARS-CoV-2 variants suggests the curve remains predictive against the Alpha, Beta, Delta, and Omicron variants where lower neutralization titers are observed (*12, 13*).

The analysis above does not allow a direct visualization or comparison of the fit of the data from breakthrough infection to the data from the vaccine comparison study. We developed a method for estimating unadjusted protection at different neutralization levels from the breakthrough infection data (see supplementary material), which also allows for inclusion of data from a third breakthrough infection study of BNT162b2 vaccinees (*5*). Figure 3 shows the data from the three breakthrough studies against the vaccine-comparison approach (normalized for the mean vaccinee titer in each study). The data from the breakthrough infection studies shows remarkable agreement with the vaccine comparison model (within the neutralization ranges where sufficient data were available for each breakthrough infection study), despite the fundamentally different data, assays, and approaches used to estimate protection curves in each study. Further, after aligning to the GMT of each vaccine group, using each of these models to predict vaccine efficacy for existing vaccines reveals good agreement between all models and the observed data, at least in the ranges where data was available to parameterize the models (Figure S1, supplement material). Together, this provides cross-validation of the protection curves, but also provides a lesson that all models should be used cautiously outside the ranges of the data over which they were developed.

**Figure 3:**
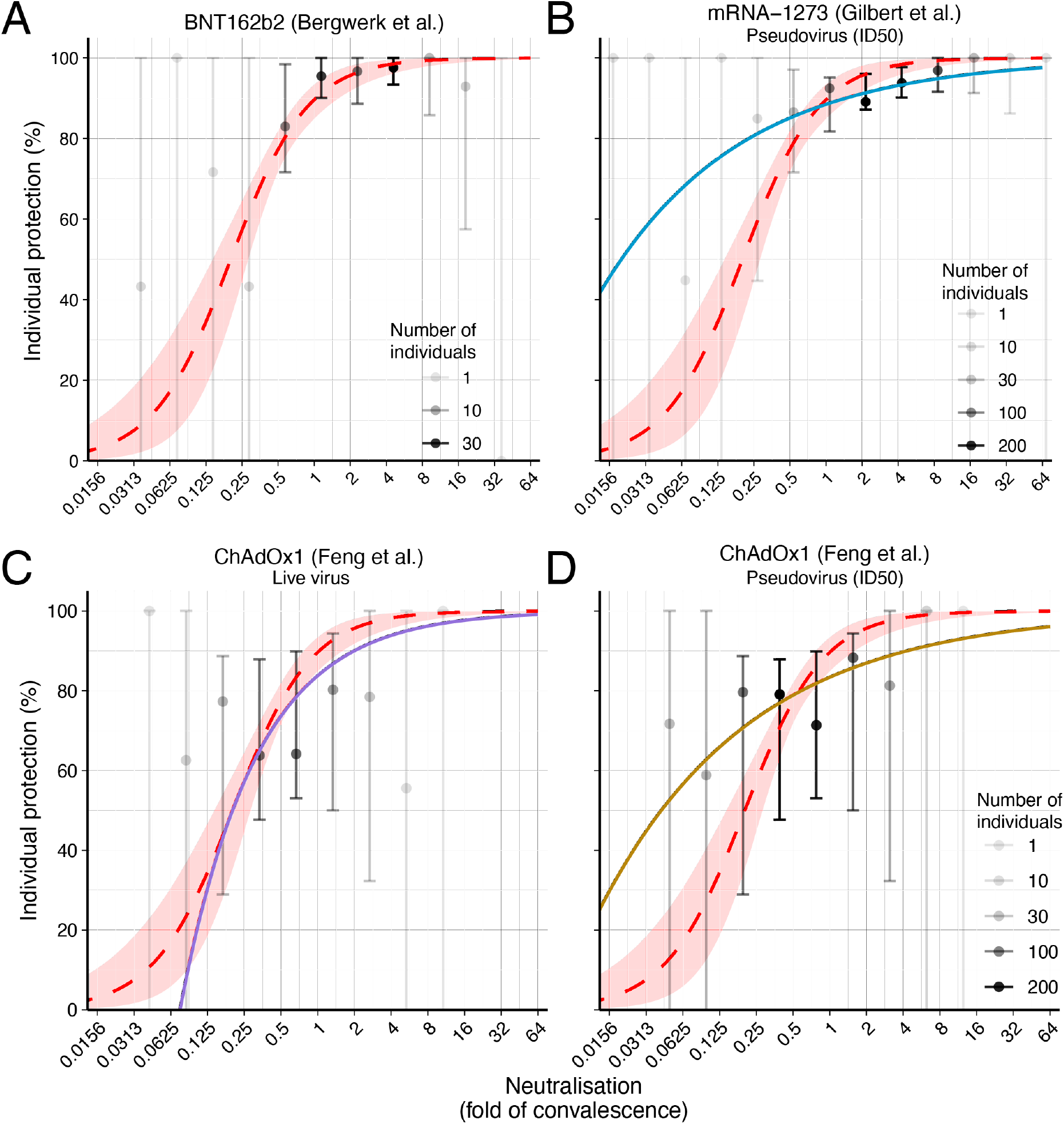
Breakthrough infection data and protection from SARS-CoV-2 infection: The relationship between neutralizing antibody titer (x-axis) and protection from symptomatic SARS-CoV-2 infection for an individual (y-axis). The protection curve derived from the vaccine-comparison model is shown (red dashed line and shading 95% confidence intervals) and is compared to the observed normalized frequencies of neutralization level (see supplement for calculations) of breakthrough infections reported in three studies (gray/black dots). Data from two mRNA vaccine studies of mRNA-1273 (A) and BNT162b2 (B), and the adenoviral vector vaccine ChAdOx1 nCoV19 (C, D) are shown. Lower opacity dots indicate fewer individuals with neutralization titers in that range. Also shown in each panel are modelled protection curves showing the relationship between individual neutralizing antibodies and protection estimated in each breakthrough infection study. Note: Breakthrough infection data of BNT162b2 vaccinees were generously supplied by the authors. The data were unavailable for the other two studies and were extracted from the original manuscript (see supplementary material). Extraction of data from Gilbert et al. were conducted manually and may be less reliable than the other studies (supplementary material).

### Using the protection curve

#### Immunobridging to predict vaccine efficacy

In vaccine development, an immune correlate to predict the efficacy of a novel vaccine without the need for large and expensive phase 3 efficacy trials would greatly accelerate the approval of novel vaccines (*14*). Similarly, for the incorporation of novel SARS-CoV-2 variant immunogens, being able to use surrogate measures to predict vaccine efficacy would be helpful. On a public health level, understanding neutralization of new variants as they arise and predicting likely population immunity to them would assist in predicting future infection risk. Finally, predicting changes in vaccine efficacy with immune waning and in cohorts with lower neutralization titers after vaccination (for example in the elderly or immunocompromised individuals) could inform the need for boosting and other immune protective strategies (*13*).

If a standardized neutralization assay were widely used, it would in principle be possible to offer a globally applicable GMT neutralization titer (‘threshold’) associated with a given level of protection, which regulators and vaccine developers could use as a target when assessing and approving vaccines, as the hemagglutination inhibition titer currently provides in influenza infection. However, the current lack of assay standardization means that no such threshold in IU can be determined that is broadly applicable across different neutralizing antibody assays. Alternatively, regulators have signaled that immunobridging studies should be carried out by comparison with existing vaccines (where efficacy has previously been determined) (*15, 16*). That is, vaccine developers need to identify a suitable existing vaccine for comparison and determine the superiority or non-inferiority margins relative to these vaccines in a randomized controlled trial (ie: how much higher neutralization titers are required to be, or how much lower the titers are permitted to be, compared to existing vaccines). Importantly, the protection curves reported so far (*2-4*), can be used to define the parameters of these non-inferiority or superiority trials. For example, using the vaccine-comparison model derived by Khoury et al (Figure 1C) we can estimate the non-inferiority or superiority margins to existing vaccines that would provide at least an 80% efficacy against ancestral virus (Table S2, Figure S2). If using mRNA-1273 or BNT162b2 as comparator vaccines, we find that a non-inferiority margin of 0.44-fold of the GMT observed in mRNA-1273 vaccinees or 0.54-fold of the GMT observed in BNT162b2 vaccinees, would provide high confidence the candidate vaccine has at least 80% efficacy (against ancestral virus). In the case of ChAdOx1 nCoV19 (with 4-week spacing of doses) as a comparator, we find that a superiority margin of 2.6-fold of GMT compared to ChAdOx1 nCoV19 vaccinees would provide similarly high confidence of >80% vaccine efficacy. Of note these margins are in strong agreement with the lower 95% CIs predicted in the breakthrough infection studies (fig. 2), which would predict that a candidate vaccine that induced 0.44-fold of the GMT for mRNA-1273 vaccinees would be expected to have an efficacy of at least 85% (for both the cID50 and cID80 assays, using the lower 95% CI), and that a margin of 2.6-fold of the mean ChAdOx1 titer would predict at least an efficacy 76% (the lower 95% CI of Feng et al. models do not reach 80% in all cases, see Lower confidence bound of the curves in Figure 2) (*3, 4*). Details of the analysis are provided in supplementary methods. The consensus of these three studies provides strong support for the use of non-inferiority and/or superiority margins in future immunobridging studies.

#### Identifying protective thresholds for individuals

A second goal for the study of ‘protective thresholds’ is to identify a ‘protective titer’ for clinical use. That is, determining a clinically relevant antibody level that can be used in a simple blood test to indicate if an individual is likely to have good protection from COVID-19. The current studies that have defined the relationship between neutralization titer and vaccine efficacy have not been designed or primarily concerned with defining such a threshold, as they only deal with estimates of vaccine efficacy at a population level. Furthermore, individual predictions from population statistics can be fraught. Unfortunately, the terminology of ‘threshold’ gives the impression that there might be an antibody level above which one is fully protected (and below which one is susceptible). However, the shape of the protection curves (figure 2) make it clear that there is a gradient of risk at different neutralization titers. Moreover, since estimates of neutralizing antibody titer have a significant standard deviation, this can lead to wide confidence intervals in estimating ‘protection’ level from a single serum sample (supplementary material). For example, using a typical duplicate-well and two-fold serum dilution neutralizing assay design (*17, 18*), an individual with a neutralizing antibody titer at exactly the level associated with 50% protection would have 95% confidence intervals on the estimated protection ranging from 15% to 85% protection (Supplementary materials), although this range will depend on the precision of a particular assay. It is worth noting that these are estimates of protection from symptomatic SARS-CoV-2 infection (which was the primary outcome of the studies analyzed), and protection against severe outcomes is achieved at lower neutralization titers (*2*). Together, this suggests that the lack of standardization between different serological assays is currently a major limitation for our ability to accurately assess individual neutralizing antibody titers and predict individual protection.

## Discussion

Predicting vaccine efficacy or a clinically useful ‘threshold’ of protection against COVID-19 would be a major advance for the field. The *in vitro* neutralization titer has been demonstrated to be well-correlated with vaccine efficacy and with an individual’s protection from symptomatic SARS-CoV-2 infection across multiple studies (*2-6, 12, 13*). We show that four studies have converged on a common prediction of the relationship between neutralization and protection against infection (within the bounds of where data were available within each study). The agreement of these studies provides strong support for the use of neutralizing antibody titers to predict the efficacy of new vaccines or vaccine efficacy against new variants (assuming the fold-drop in neutralization titer for the variant can be estimated). While neutralizing antibody levels are a clear correlate of protection, identifying a ‘protective threshold’ applicable to a serological test is more challenging. In part this is because no such threshold exists, but rather there is a gradient of vaccine efficacy that increases with neutralization. Further, the diversity of assays used to measure neutralization, the difficulty in translating neutralization levels between assays, the constant emergence of new and more escaped variants as well as the uncertainties in estimating individual neutralization titers present significant challenges to defining a particular ‘threshold’ at which an individual’s neutralization titer might be deemed to provide ‘high protection’ from COVID-19.

An additional major challenge is in adapting assays (and protection curves) to deal with neutralization of current and future SARS-CoV-2 variants. The studies discussed in this analysis primarily deal with neutralization of and protection from the ancestral (Wuhan-like) SARS-CoV-2 strain since the breakthrough infection data and vaccine efficacy data in most studies was from the Phase 3 clinical trials (*2-4*), which studied infection within the first few months after vaccination, and which mainly occurred before variants of concern had a major foothold (with the exception of reference (*5*), which occurred in the Alpha-dominant period). We have recently shown that the vaccine-comparison model calibrated on data for ancestral SARS-CoV-2 infections can also be used to predict vaccine effectiveness against SARS-CoV-2 variants and after boosting if one adjusts for the drop in neutralization titers to the variants and rise in neutralization after boosting (*13, 19, 20*). However, the need to standardize neutralization assays for SARS-CoV-2 variants presents an ongoing challenge.

*In vitro* neutralizing antibody titers against SARS-CoV-2 present a clear correlate of protection from symptomatic SARS-CoV-2 infection. Studies of passive administration of neutralizing monoclonal antibodies in animals and humans support that this is a mechanistic correlate of protection (*21-23*). Indeed, a recent study comparing protective titers in prophylactic and therapeutic studies suggests that the protective titers may be very similar (*24*). Neutralizing antibody levels are also correlated with protection from severe SARS-CoV-2 infection (*2*). However, it is likely that other immune responses may also play a significant role in protecting from progression from symptomatic to severe SARS-CoV-2 infection and further work is required to better understand the determinants of severe COVID-19 outcomes.

The agreement across multiple studies of the relationship between neutralizing antibodies and efficacy against COVID-19 has important implications for future vaccine use, population immunity and reducing the global impact of the COVID-19 pandemic.

## Data Availability

All data and code will be made available on GitHub upon publication.

## Glossary

Protection curve: The relationship between the measured immune response of a vaccine in a subgroup of individuals, and the level of protection from symptomatic infection provided by the vaccine in that subgroup compared to placebo (protection = vaccine efficacy).
Threshold of protection: The level of immune response required to provide a specified level of protection (vaccine efficacy) from COVID-19. The 50% protective threshold is commonly reported.
Fold-of-convalescent scale: An attempt to compare between different assays by normalizing titers to that of convalescent subjects in the same assay. Requires convalescent subjects to have similar infection histories for accurate comparison.
International Units per milliliter (IU/ml): A neutralization titer (or mean neutralization titer) calibrated to a WHO international standard and reported in IU/ml.

## Acknowledgements

The authors wish to thank Moriah Bergwerk, Tal Gonen, Gili Regev-Yoshay and colleagues for supplying the raw data from their study of neutralization titers in BNT162b2 vaccinated healthcare workers.

## Ethics statement

This work was approved under the UNSW Sydney Human Research Ethics Committee (approval HC200242).

## Authorship Statement

DC, MS, JAT, DSK and MPD contributed to conceptualization, supervision, and resources. DC, MS, AR, TES, DSK and MPD contributed to data curation, methodology, formal analysis, and visualization. All authors contributed to the data collection and writing and reviewed and approved the final report. All authors had full access to all the data in the study and had final responsibility for the decision to submit for publication.

## Data Availability Statement

All data and code will be made available on GitHub upon publication.

## References

1. J. Huddleston, J. R. Barnes, T. Rowe, X. Xu, R. Kondor, D. E. Wentworth, L. Whittaker, B. Ermetal, R. S. Daniels, J. W. McCauley, S. Fujisaki, K. Nakamura, N. Kishida, S. Watanabe, H. Hasegawa, I. Barr, K. Subbarao, P. Barrat-Charlaix, R. A. Neher, T. Bedford, Integrating genotypes and phenotypes improves long-term forecasts of seasonal influenza A/H3N2 evolution. Elife 9, (2020).

2. D. S. Khoury, D. Cromer, A. Reynaldi, T. E. Schlub, A. K. Wheatley, J. A. Juno, K. Subbarao, S. J. Kent, J. A. Triccas, M. P. Davenport, Neutralizing antibody levels are highly predictive of immune protection from symptomatic SARS-CoV-2 infection. Nat Med 27, 1205–1211 (2021).

3. S. Feng, D. J. Phillips, T. White, H. Sayal, P. K. Aley, S. Bibi, C. Dold, M. Fuskova, S. C. Gilbert, I. Hirsch, H. E. Humphries, B. Jepson, E. J. Kelly, E. Plested, K. Shoemaker, K. M. Thomas, J. Vekemans, T. L. Villafana, T. Lambe, A. J. Pollard, M. Voysey, C. V. T. G. Oxford, Correlates of protection against symptomatic and asymptomatic SARS-CoV-2 infection. Nat Med, (2021).

4. P. B. Gilbert, D. C. Montefiori, A. B. McDermott, Y. Fong, D. Benkeser, W. Deng, H. Zhou, C. R. Houchens, K. Martins, L. Jayashankar, F. Castellino, B. Flach, B. C. Lin, S. O’Connell, C. McDanal, A. Eaton, M. Sarzotti-Kelsoe, Y. Lu, C. Yu, B. Borate, L. W. P. v. d. Laan, N. S. Hejazi, C. Huynh, J. Miller, H. M. E. Sahly, L. R. Baden, M. Baron, L. D. L. Cruz, C. Gay, S. Kalams, C. F. Kelley, M. P. Andrasik, J. G. Kublin, L. Corey, K. M. Neuzil, L. N. Carpp, R. Pajon, D. Follmann, R. O. Donis, R. A. Koup, Immune correlates analysis of the mRNA-1273 COVID-19 vaccine efficacy clinical trial. Science 375, 43–50 (2022).

5. M. Bergwerk, T. Gonen, Y. Lustig, S. Amit, M. Lipsitch, C. Cohen, M. Mandelboim, E. G. Levin, C. Rubin, V. Indenbaum, I. Tal, M. Zavitan, N. Zuckerman, A. Bar-Chaim, Y. Kreiss, G. Regev-Yochay, Covid-19 Breakthrough Infections in Vaccinated Health Care Workers. N Engl J Med 385, 1474–1484 (2021).

6. K. A. Earle, D. M. Ambrosino, A. Fiore-Gartland, D. Goldblatt, P. B. Gilbert, G. R. Siber, P. Dull, S. A. Plotkin, Evidence for antibody as a protective correlate for COVID-19 vaccines. Vaccine 39, 4423–4428 (2021).

7. W. H. Organization, Establishment of the WHO International Standard and Reference Panel for anti-SARS-CoV-2 antibody. Expert Committee on Biological Standardization, Geneva, 9–10 (2020).

8. P. M. Folegatti, K. J. Ewer, P. K. Aley, B. Angus, S. Becker, S. Belij-Rammerstorfer, D. Bellamy, S. Bibi, M. Bittaye, E. A. Clutterbuck, C. Dold, S. N. Faust, A. Finn, A. L. Flaxman, B. Hallis, P. Heath, D. Jenkin, R. Lazarus, R. Makinson, A. M. Minassian, K. M. Pollock, M. Ramasamy, H. Robinson, M. Snape, R. Tarrant, M. Voysey, C. Green, A. D. Douglas, A. V. S. Hill, T. Lambe, S. C. Gilbert, A. J. Pollard, C. V. T. G. Oxford, Safety and immunogenicity of the ChAdOx1 nCoV-19 vaccine against SARS-CoV-2: a preliminary report of a phase 1/2, single-blind, randomised controlled trial. Lancet 396, 467–478 (2020).

9. D. S. Khoury, A. K. Wheatley, M. D. Ramuta, A. Reynaldi, D. Cromer, K. Subbarao, D. H. O’Connor, S. J. Kent, M. P. Davenport, Measuring immunity to SARS-CoV-2 infection: comparing assays and animal models. Nat Rev Immunol 20, 727–738 (2020).

10. L. A. Jackson, E. J. Anderson, N. G. Rouphael, P. C. Roberts, M. Makhene, R. N. Coler, M. P. McCullough, J. D. Chappell, M. R. Denison, L. J. Stevens, A. J. Pruijssers, A. McDermott, B. Flach, N. A. Doria-Rose, K. S. Corbett, K. M. Morabito, S. O’Dell, S. D. Schmidt, P. A. Swanson, 2nd, M. Padilla, J. R. Mascola, K. M. Neuzil, H. Bennett, W. Sun, E. Peters, M. Makowski, J. Albert, K. Cross, W. Buchanan, R. Pikaart-Tautges, J. E. Ledgerwood, B. S. Graham, J. H. Beigel, R. N. A. S. G. m, An mRNA Vaccine against SARS-CoV-2 - Preliminary Report. N Engl J Med 383, 1920–1931 (2020).

11. L. R. Baden, H. M. El Sahly, B. Essink, K. Kotloff, S. Frey, R. Novak, D. Diemert, S. A. Spector, N. Rouphael, C. B. Creech, J. McGettigan, S. Khetan, N. Segall, J. Solis, A. Brosz, C. Fierro, H. Schwartz, K. Neuzil, L. Corey, P. Gilbert, H. Janes, D. Follmann, M. Marovich, J. Mascola, L. Polakowski, J. Ledgerwood, B. S. Graham, H. Bennett, R. Pajon, C. Knightly, B. Leav, W. Deng, H. Zhou, S. Han, M. Ivarsson, J. Miller, T. Zaks, C. S. Group, Efficacy and Safety of the mRNA-1273 SARS-CoV-2 Vaccine. N Engl J Med 384, 403–416 (2021).

12. D. S. Khoury, M. Steain, J. A. Triccas, A. Sigal, M. P. Davenport, D. Cromer, A meta-analysis of Early Results to predict Vaccine efficacy against Omicron. medRxiv, 2021.2012.2013.21267748 (2021).

13. D. Cromer, M. Steain, A. Reynaldi, T. E. Schlub, A. K. Wheatley, J. A. Juno, S. J. Kent, J. A. Triccas, D. S. Khoury, M. P. Davenport, Neutralising antibody titres as predictors of protection against SARS-CoV-2 variants and the impact of boosting: a meta-analysis. The Lancet Microbe 3, e52–61 (2021).

14. E. B. Walter, K. R. Talaat, C. Sabharwal, A. Gurtman, S. Lockhart, G. C. Paulsen, E. D. Barnett, F. M. Munoz, Y. Maldonado, B. A. Pahud, J. B. Domachowske, E. A. F. Simoes, U. N. Sarwar, N. Kitchin, L. Cunliffe, P. Rojo, E. Kuchar, M. Ramet, I. Munjal, J. L. Perez, R. W. Frenck, Jr., E. Lagkadinou, K. A. Swanson, H. Ma, X. Xu, K. Koury, S. Mather, T. J. Belanger, D. Cooper, O. Tureci, P. R. Dormitzer, U. Sahin, K. U. Jansen, W. C. Gruber, C. C. T. Group, Evaluation of the BNT162b2 Covid-19 Vaccine in Children 5 to 11 Years of Age. N Engl J Med 386, 35–46 (2022).

15. Medicines & Healthcare products Regulatory Agency, Access Consortium: Alignment with ICMRA consensus on immunobridging for authorising new COVID-19 vaccines. https://www.gov.uk/government/publications/access-consortium-alignment-with-icmra-consensus-on-immunobridging-for-authorising-new-covid-19-vaccines/access-consortium-alignment-with-icmra-consensus-on-immunobridging-for-authorising-new-covid-19-vaccines, xAccessed: 8 April 2022 (2021).

16. Medicines & Healthcare products Regulatory Agency, Guidance on strain changes in authorised COVID-19 vaccines. https://www.gov.uk/government/publications/access-consortium-guidance-on-strain-changes-in-authorised-covid-19-vaccines/guidance-on-strain-changes-in-authorised-covid-19-vaccines, xAccessed: 8 April 2022 (2021).

17. J. A. Juno, H. X. Tan, W. S. Lee, A. Reynaldi, H. G. Kelly, K. Wragg, R. Esterbauer, H. E. Kent, C. J. Batten, F. L. Mordant, N. A. Gherardin, P. Pymm, M. H. Dietrich, N. E. Scott, W. H. Tham, D. I. Godfrey, K. Subbarao, M. P. Davenport, S. J. Kent, A. K. Wheatley, Humoral and circulating follicular helper T cell responses in recovered patients with COVID-19. Nat Med 26, 1428–1434 (2020).

18. A. K. Wheatley, J. A. Juno, J. J. Wang, K. J. Selva, A. Reynaldi, H. X. Tan, W. S. Lee, K. M. Wragg, H. G. Kelly, R. Esterbauer, S. K. Davis, H. E. Kent, F. L. Mordant, T. E. Schlub, D. L. Gordon, D. S. Khoury, K. Subbarao, D. Cromer, T. P. Gordon, A. W. Chung, M. P. Davenport, S. J. Kent, Evolution of immune responses to SARS-CoV-2 in mild-moderate COVID-19. Nat Commun 12, 1162 (2021).

19. S. Cele, L. Jackson, D. S. Khoury, K. Khan, T. Moyo-Gwete, H. Tegally, J. E. San, D. Cromer, C. Scheepers, D. G. Amoako, F. Karim, M. Bernstein, G. Lustig, D. Archary, M. Smith, Y. Ganga, Z. Jule, K. Reedoy, S. H. Hwa, J. Giandhari, J. M. Blackburn, B. I. Gosnell, S. S. Abdool Karim, W. Hanekom, S. A. Ngs, C.-K. Team, A. von Gottberg, J. N. Bhiman, R. J. Lessells, M. S. Moosa, M. P. Davenport, T. de Oliveira, P. L. Moore, A. Sigal, Omicron extensively but incompletely escapes Pfizer BNT162b2 neutralization. Nature 602, 654–656 (2022).

20. D. Cromer, A. Reynaldi, M. Steain, J. A. Triccas, M. P. Davenport, D. S. Khoury, Relating in vitro neutralisation level and protection in the CVnCoV (CUREVAC) trial. Clin Infect Dis In Press, ciac075 (2022).

21. S. A. Plotkin, Vaccines: correlates of vaccine-induced immunity. Clin Infect Dis 47, 401–409 (2008).

22. M. P. O’Brien, E. Forleo-Neto, B. J. Musser, F. Isa, K. C. Chan, N. Sarkar, K. J. Bar, R. V. Barnabas, D. H. Barouch, M. S. Cohen, C. B. Hurt, D. R. Burwen, M. A. Marovich, P. Hou, I. Heirman, J. D. Davis, K. C. Turner, D. Ramesh, A. Mahmood, A. T. Hooper, J. D. Hamilton, Y. Kim, L. A. Purcell, A. Baum, C. A. Kyratsous, J. Krainson, R. Perez-Perez, R. Mohseni, B. Kowal, A. T. DiCioccio, N. Stahl, L. Lipsich, N. Braunstein, G. Herman, G. D. Yancopoulos, D. M. Weinreich T. Covid-19 Phase 3 Prevention Trial, Subcutaneous REGEN-COV Antibody Combination to Prevent Covid-19. N Engl J Med 385, 1184–1195 (2021).

23. M. S. Cohen, A. Nirula, M. J. Mulligan, R. M. Novak, M. Marovich, C. Yen, A. Stemer, S. M. Mayer, D. Wohl, B. Brengle, B. T. Montague, I. Frank, R. J. McCulloh, C. J. Fichtenbaum, B. Lipson, N. Gabra, J. A. Ramirez, C. Thai, W. Chege, M. M. Gomez Lorenzo, N. Sista, J. Farrior, M. E. Clement, E. R. Brown, K. L. Custer, J. Van Naarden, A. C. Adams, A. E. Schade, M. C. Dabora, J. Knorr, K. L. Price, J. Sabo, J. L. Tuttle, P. Klekotka, L. Shen, D. M. Skovronsky, B.-. Investigators, Effect of Bamlanivimab vs Placebo on Incidence of COVID-19 Among Residents and Staff of Skilled Nursing and Assisted Living Facilities: A Randomized Clinical Trial. JAMA 326, 46–55 (2021).

24. E. Stadler, K. L. Chai, T. E. Schlub, D. Cromer, M. N. Polizzotto, S. J. Kent, N. Skoetz, L. Estcourt, Z. K. McQuilten, E. M. Wood, D. S. Khoury, M. P. Davenport, Determinants of passive antibody effectiveness in SARS-CoV-2 infection. medRxiv, 2022.2003.2021.22272672 (2022).

