## Supplementary Material for "Correlates of protection, thresholds of protection, and immunobridging in SARS-CoV-2 infection"

**Supplementary Methods and Analysis**

**Conversion of Khoury et al., fold-convalescence scale to WHO international units (IU)**
The geometric mean neutralization titer in convalescent subjects at ~30 day post symptom onset in a cohort of 56 individuals described in (*1*) (and used in ref (*2*)) was measured (considering censoring, (*1*)) as 55.7. In the same laboratory, the same microneutralization assay was performed on the international standard 20/130, reported to equate to 1300 IU/ml (95% CI: 981-1719)(*3*). The assay was performed on the standard 29 times, with a geometric mean neutralization titer of 272 (95% CI: 234-316). Therefore, the geometric mean neutralization titer of convalescent subjects is given by $55.7 \times 1300/272 = 266$ IU/ml, and the 50% protective titer calculated in(*1*) is $20.11\% \times 266/100 = 53.5$ IU/ml.

In Khoury et al.(*1*), using data from Phase 1/2 trials, the mRNA-1273 and ChAdOx1 nCoV-19 vaccines were estimated to have geometric neutralization titers of 4.1-fold and 0.54-fold of the geometric mean titers of convalescent individuals, respectively. Therefore, in WHO international units these geometric mean neutralization titers for each vaccine equate to $266 \times 4.1 = 1099$ IU/ml for mRNA-1273 and $266 \times 0.54 = 144$ IU/mL for ChAdOx1 nCoV-19.

**The role of differing neutralization titers assays in the different threshold of protection reported in the literature**
Three these studies have estimated the level of protection associated with particular antibody levels and reported a 'threshold' antibody level required for 50% or 70% protection from symptomatic SARS-CoV-2 infection (*1, 4, 5*). There was not agreement between all of these studies, with some of the three studies yielding quite different estimates of the protection curves and thresholds of protection from symptomatic infection, especially when different assays used to assess neutralizing antibody titers. This is a critical issue for the field to reconcile in order to move forward with a defined correlate of protection. Khoury et al. used a meta-analysis of Phase 1/2 and Phase 3 vaccine trials (vaccine comparison approach, Figure 1) and estimated that 50% vaccine efficacy was achieved with a neutralization titer of 54 IU/ml (in a live virus neutralization assay). More recently, two studies used data from subjects with 'breakthrough infection' after antibody responses were measured following two doses of mRNA 1273 (n= 36) or ChAdOx1 nCoV19 (n= 47) vaccination to model the protection curve (Figure 1D-F). Across these studies both pseudovirus and live SARS-CoV-2 neutralization assays were used to measure neutralizing antibodies. Interestingly, the estimated curves using data from the same pseudoviral neutralization IC50 assay yielded similar 70% protective threshold of 4 IU/ml and 8 IU/ml, respectively (*4, 5*) (Figure S3). However, Feng et al. used both a pseudovirus neutralization assay and a live SARS-CoV-2 neutralization assay, and showed that the latter yielded a 70% protective titer that was >4-fold higher than the pseudovirus assay (in international units)(*4*). Given the large variance in neutralizing antibody reports for notionally similar groups of vaccinated individuals when different assays are employed, even after conversion to IU, it is perhaps unsurprising that similar assays yield more similar results once converted to IU, but it would appear that the different assays contribute directly to different estimated titers required for the same level of protection (even within the same study using the same breakthrough infections).

**Normalizing neutralization titers between studies.**

To normalize neutralization titers between the vaccine comparison study(*1*), and the breakthrough infection studies (*4-6*) we assumed that the geometric mean (all references to mean of neutralization titers in this text means geometric mean) neutralization titer of individuals vaccinated with a particular vaccine should be equivalent across studies. For example, when comparing neutralization data from the Khoury et al. and Gilbert et al. studies, we noted that in the Gilbert et al. study only mRNA-1273 vaccinees were considered, and in the uninfected mRNA-1273 vaccines the mean neutralization titer reported for the ID50 assay was 247 IU/ml. Similarly, in the Khoury et al. study the mean neutralization titer for mRNA-1273 vaccinees was taken from (*7*) and was estimated as 4.1-fold of the mean of convalescent individuals analysed in the same study. Further, we assume that neutralization titers across the different assays in the different studies will vary colinearly. Thus, to align all ID50 measurements and curves in the Gilbert et al. and Khoury et al. studies we divided all the titers by the GMT of 247 IU/ml (for Gilbert et al.) and 4.1-fold (for Khoury et al.). In addition, for visualization, we place the ID50 data from the Gilbert et al. study on the fold-of-convalescence scale reported in Khoury et al., by multiplying these aligned curves by 4.1. In the case of Feng et al. and Bergwerk et al., the same approach was used but using the geometric mean neutralization titer for ChAdOx1 nCoV-19 and BNT162b2, respectively (Feng et al. reported the median neutralization titer which was used in place of the geometric mean).

It is important to note that the above normalization assumes that the mean neutralization titer in the Phase 1/2 studies of each of these vaccines(*7-9*) (used in the Khoury et al.) is approximately equivalent to the mean neutralization titer for the corresponding vaccine in the breakthrough-infection studies (*4-6*). Of note is that vaccination schedules were not equivalent for ChAdOx1 nCoV-19 vaccinees in the Phase 1/2 trial (4 week schedule) and the Feng et al. study (a mixture of prime-boost schedules of between <6 week and >12 week), where dose spacing was altered from the intended schedule due to supply issues(*10*). The mixture of dosing schedules is likely to make the mean neutralization titers differ between the studies. We assessed the impact of this difference using data reported in the meta-analysis of these neutralization results from a large number of the Phase 3 trial participants(*11*). Performing a weighted average of the neutralization titers of the whole population from this meta-analysis (which resembles the cohort in the Feng et al. study) we found the mean neutralization titer was only 1.40-fold higher than the average of vaccinees with doses spaced by <6 weeks (*11*) (i.e. the <6 week spacing is comparable with the Phase 1/2 trial cohort dosing regimen (*9*)). This demonstrated that differences in dosing schedules will only have a minor impact on the overall normalization of the Feng et al. study with the Khoury et al. study. Thus, in the main text we use the raw mean neutralization titers from the Phase 1/2 and Phase 3 trials for simplicity.

**Extracting the model relating neutralization and protection**

In the Gilbert et al.(*5*) and Feng et al.(*4*) studies a mathematical model was used to estimate the relationship between neutralization titer and protection. This relationship was modelled for two different neutralization assays per study and the final model and confidence intervals were extracted from figure 4 in (*4*) and figures 4 and S23 in (*5*) using the WebPlotDigitizer online application (https://automeris.io/WebPlotDigitizer/). These extracted models were normalized to a fold-of-convalescence scale (as described above) and plotted in figure 2 against the fitted model reported in Khoury et al (*1*).

**Data on breakthrough infection**

Data was requested from the authors of(*4-6*). Raw data were provided by the authors for(*6*). Data were unavailable for the other two studies, and therefore were extracted from the published work. Data were extracted using Adobe Illustrator (by saving figures in an SVG format and using a text editor to extract coordinates of datapoints from the vector graphic images contained in the publication) from Extended data figure 2 in(*4*), and the WebPlotDigitizer online application ([https://automeris.io/WebPlotDigitizer/](https://automeris.io/WebPlotDigitizer/)) from figure S10 in(*5*). The neutralization titres of the uninfected vaccinated groups and the symptomatic breakthrough infections groups were extracted. Neutralization data on control and breakthrough infections from(*6*) were provided by the authors.

**Calculating the unadjusted protection curve from breakthrough infection data to compare with the fitted models**

In the three breakthrough infection studies reported (*4-6*), two groups of individuals are considered, vaccinated individuals with breakthrough infection and vaccinated individuals without breakthrough infection. Importantly, their uninfected status is not necessarily due to vaccine protection but in many cases will reflect simply that those individuals were not exposed to COVID-19. Hence the Gilbert et al., and Feng et al., studies used an additional risk model to characterize and account for the unvaccinated placebo arm's rate of symptomatic SARS-CoV-2 infection. A challenge with the breakthrough infection studies is visualizing the data and the risk of symptomatic infection at different neutralization titers. Here we use the neutralization data reported in these three breakthrough infection studies to visualize an unadjusted protection curve (figure 3). These unadjusted protection curves were calculated by assuming the following:

1. Control data on the neutralization titers of individuals who received the vaccine but were not infected, provides a reasonable approximation of the distribution of neutralization titers in individuals who were vaccinated (independent of breakthrough infections). Despite control populations inherently excluding individuals who experienced breakthrough infections, as the proportion of breakthrough infections is small (<7%), any bias resulting from this assumption is likely to be small.
2. Let $i$ and $e$ be the events of an individual being infected, and an individual being exposed to SARS-Cov-2 (in a manner that would cause infection in an unvaccinated individual) respectively. $P(i)$ and $P(e)$ denotes the probabilities of these events. Then

$$P(i) = P(e)(1 - E)$$

where E is the vaccine efficacy. Therefore,

$$\frac{P(i)}{P(e)} = 1 - E.$$

3. The probability of exposure is independent of neutralization titer, i.e.

$$P(e \cap n) = P(e)P(n)$$

Given the above, we are primarily concerned with calculating the probability that an individual becomes infected given that they are exposed and have a given neutralization titer ($n$). Denote an individual's risk of becoming infected given that they are exposed and have a neutralization titer ($n$) as, $P(i|e \cap n)$, therefore:

$$P(i|e \cap n) = \frac{P(i \cap e \cap n)}{P(e \cap n)} = \frac{P(i \cap e \cap n)}{P(e)P(n)} \quad (3)$$

We note, using conditional probability, and noting that the probability an individual was exposed if they were infected is 1, i.e. $P(e \cap i) = P(i)$:

$$P(n|e \cap i) = \frac{P(i \cap e \cap n)}{P(e \cap i)} = \frac{P(i \cap e \cap n)}{P(i)} \quad (4)$$

Combining equations 3 and 4 above, and using assumption 2, it follows that:

$$P(i|e \cap n) = \frac{P(i)P(n|e \cap i)}{P(e)P(n)} = (1 - E)\frac{P(n|i)}{P(n)} \quad (5)$$

Therefore, the probability that an individual who is exposed becomes infected, given that they have a neutralization titer ($n$), is equal to the ratio of the probability that a breakthrough infection (Case) has a neutralization titer ($n$), divided by the probability an uninfected vaccinee has a neutralization titer n, and this is multiplied by 1 minus the vaccine efficacy. Using the neutralization data reported in each of the three breakthrough infection studies, we performed binning of neutralization data into bins spaced by 2-fold titers, and for each neutralization bin (corresponding to a neutralization titer $n$), we calculated the fraction of individuals with each neutralization titer $n$ who had a breakthrough infection (i.e. $P(n|i)$) and the fraction of individuals in the control vaccinee group (i.e. $P(n)$). Since the breakthrough infection data for Gilbert et al. and Feng et al., were taken from randomized placebo-controlled Phase 3 trials, the vaccine efficacy, $E$, within these specific populations was determined in the papers and risk model themselves (for Gilbert et al., efficacy was reported in figure 4 of the paper and for Feng et al. the overall efficacy was back-calculated using the model reported in this study using the approach described in *Immunobridging: Predicting the efficacy of another vaccine*), and for the Bergwerk et al. study an observational study of the vaccine efficacy from the same region with overlapping calendar time was used for an estimate of the vaccine efficacy in that setting (*12*) (table S5). These unadjusted protection curves were normalized to the fold-of-convalescence scale in the same way as described earlier, to generate the figure 3.

The confidence intervals of these unadjusted estimates of protection were determined by parametric bootstrapping of the neutralization titers. That is, we first fitted the (extracted or provided) neutralization data for individuals with breakthrough infection ("cases", assume $n_{case}$ number of individuals) and uninfected vaccinated individuals ("uninfected-vaccinated", assume $n_{uninfected}$ number of individuals) with a normal distribution using censoring regression(*1*). These fitted distributions of the neutralization titers for cases and uninfected-vaccinated from each study were then sampled randomly $n_{case}$ and $n_{uninfected}$ times, respectively, and the resulting data were used to recalculate the unadjusted protection of individuals within each 2-fold range of neutralization titers (as describe above). This was repeated 10,000 times for each study, and the 2.5$^{th}$ and 97.5$^{th}$ percentiles of the 10,000 estimates of the unadjusted protection were used to estimate the confidence intervals. Some iterations produced missing values for the vaccine efficacy estimate because by random sampling some ranges of neutralization titers had no uninfected-vaccinated individuals – in this case we did two things, we excluded the missing iterations from the calculation of the confidence intervals and we also set these missing values to an extreme estimate of 1 or 0 (i.e. 0% protection or 100% protection) and recalculated the 95% CIs. We then took the maximum of these two approaches as the Upper bound of the 95% CI, and the minimum of these two approaches as the Lower bound of the 95% CI.

**Estimating the standard deviation of neutralization titers**

For the Feng et al. study, raw neutralization titers could be precisely extracted and in this case censoring regression was used to fit a normal distribution to the data to estimate the standard deviation of neutralization titers from both assays reported in that study (*4*). However, for the Gilbert et al. study, the raw data were not available and so the standard deviation of the neutralization titers were calculated from the confidence interval reported for the means in table 1 of(*5*). This was performed as follows:

$$SD_1 = \left(log10(M) - log10(L)\right) \times \frac{\sqrt{1005}}{1.96}$$

$$SD_2 = \left(log10(U) - log10(M)\right) \times \frac{\sqrt{1005}}{1.96}$$

$$\sigma = \frac{SD_1 + SD_2}{2}$$

where, $M$ is the geometric mean titer of the uninfected vaccinated population, $L$ and $U$ are the lower and upper bounds of the 95% CI of the mean neutralization titer for the uninfected vaccinated population and $SD_1$ and $SD_2$ are two estimates of the SD from the lower and upper bounds of the 95% CI, respectively. Note that neutralization titers for 1005 uninfected individuals vaccinated with mRNA-1273 were reported in this study.

We found that the standard deviation of the neutralization titers were close (Table S3, range of 0.41-0.47, across the different neutralization markers measured in the different two studies) to the standard deviation of neutralization titers estimated by Khoury et al. (*1*), when all neutralization titers from all phase 1/2 studies were pooled (0.46).

**Immunobridging: Predicting the efficacy of another vaccine (generating Figure S1)**

Using the models from Gilbert et al. and Feng et al., that estimate the relationship between neutralization and an individual's protection from symptomatic SARS-CoV-2 infection it is possible to predict the efficacy of another vaccine if one knows the distribution of neutralization titers induced by that vaccine. To calculate the predicted vaccine efficacy for a vaccine $V$, using each of the published models, one approach is based on the following formula:

$$PE(V) = \int_m^M V(n)P(n)\,dn$$

where, $V(n)$ is the empirical probability density of $\log_{10}$ neutralization titers induced by the vaccine $V$, and $P(n)$ is the estimated protection (i.e. equivalent to the "vaccine efficacy" or "controlled vaccine efficacy" curves as reported in Feng et al. and Gilbert et al., respectively) for a given $\log_{10}$ neutralization titer as reported in the models published in (*4, 5*). $M$ and $m$ are the maximum and minimum $\log_{10}$ neutralization titers. The estimated protection (vaccine efficacy) level given a $\log_{10}$ neutralization titer ($n$) is given in the Gilbert et al. and Feng et al. studies by combining multiple methodologies, including inverse probability weightings, a Cox regression model (with an estimate of the baseline hazard) (curves in figure 4,S23 and figure 4 of those studies, respectively). However, under the causal assumptions of(*13*) (no unmeasured confounders and positivity), with a simplified approach that does not control for baseline covariates, the combined functional form of this model can be written as

$$P(n) = 1 - c(1 - e^{-ae^{-b\,n}}). \qquad (1)$$

We estimated values for *a, b* and *c* by fitting the curve in equation 1 to points extracted from figure 4,S23 and figure 4 of (*5*) and (*4*), respectively. Fitting was performed using a standard least squares approach, to the natural log of the extracted values (extraction described above) (fig. S4 and table S4).

For any vaccine that induces approximately normally distributed neutralization titers (mean $\mu$, standard deviation $\sigma_s$) we can then estimate the predicted vaccine efficacy (PE($\mu$)) using the Gilbert et al. and Feng et al. models of protection (figure S4) as:

$$PE(\mu) = \int_{-\infty}^{\infty} N(n|\mu, \sigma_s)\left(1 - c(1 - e^{-ae^{-b\,n}})\right)\,dn, \qquad (2)$$

where $N$ is the probability density function of a normal distribution with mean $\mu$ and standard deviation $\sigma_s$. Note that the neutralization titers in Feng et al., and Gilbert et al. appear approximately normally distributed (Figure S5).

Using equation 2 and the estimated parameters in Table S3 and S4, we calculated the efficacy of other vaccines that would be predicted by the models in the Gilbert et al. and Feng et al. studies (Figure S1).

**Estimating non-inferiority or superiority margins that will give high confidence of at least 80% efficacy for a candidate vaccine**

It is useful for regulators to be able to define minimum criteria for vaccine developers to meet in order to define an effective new agent based on neutralizing antibodies. However, given assay variability it is not possible to define a particular neutralizing antibody titer that should be achieved by a new vaccine in order for it to have a certain vaccine efficacy. Instead, direct comparison of a new candidate vaccine against an existing comparator vaccine in a non-inferiority or superiority trial will be a more robust approach. Here we estimate what difference in geometric mean neutralization titers between a candidate

vaccine and an existing vaccine is acceptable/necessary in order for there to be a high confidence the candidate vaccine has at least 80% efficacy. This analysis can also be adjusted to report superiority / non-inferiority margins for other efficacy thresholds.
Using the model reported by Khoury et al. (*1*), we can predict vaccine efficacy for a given geometric mean neutralization titer normalized to the mean of a convalescent panel, but there are a number of sources of uncertainty. In particular, the mean neutralization titer of the comparator vaccine has uncertainty (we consider three comparator vaccines here BNT162b2, mRNA-1273 and ChAdOx1 nCoV-19), and the model itself had uncertainty (see 95% CI's in figure 2). Thus, for a given reference vaccine with a mean ($\log_{10}$) neutralization titer $m$, and a given fold-change in the geometric mean neutralization titer of a new candidate vaccine compared with that reference vaccine ($10^d$), we compute the lower 95% confidence bound of the estimated vaccine efficacy for the new vaccine with, using the model reported by (*1*):

$$VE(m) = \int_{-\infty}^{\infty} N(n|m+d,\sigma) \frac{1}{1+e^{-k(n-n_{50})}} dn$$

where, $m$ is the $\log_{10}$ of the neutralization titer (on a fold of convalescence scale) of the reference vaccine, $n_{50}$ is the ($\log_{10}$) neutralization titer estimated to give 50% VE, $k$ is the slope parameter relating neutralization and vaccine efficacy, and $N$ is the probability density function of a normal distribution representing the distribution in neutralization titers induced by a given vaccine mean ($\log_{10}$) neutralization titer of $m$ and standard deviation of ($\log_{10}$) neutralization titers of $\sigma$. To compute the lower 95% confidence bound from this model, we use parametric bootstrapping (as in (*14*)). Briefly, we estimate the vaccine efficacy using the above model 50,000 times, after sampling the model parameters at random from a normal distribution to capture the uncertainty in these parameters. The normal distributions used for randomly sampling the parameters are assumed to have means given by the parameter estimates and standard deviations given by the standard error (and covariance matrix for jointly distributed parameters) of the estimated parameters $n_{50}$, $\log(k)$ and $\sigma$ obtained during model fitting in the original study (*1*). Given that the mean neutralization titer of the reference vaccine ($m$) also contains uncertainty, we similarly, draw this parameter randomly using the standard error in $m$ estimated in the original study (*1*) (horizontal confidence bands, in Figure S2). Of the 50,000 repeated estimates we then take the lower 5$^{th}$ percentile of these bootstrapped predictions to estimate the one-tailed lower 95% confidence interval of candidate vaccine efficacy given the fixed change in neutralization titer (shaded region in Figure S2). We then find the change in neutralization titer of the novel agent compared to the existing comparator vaccine that will provide a lower bound on the vaccine efficacy confidence interval of 80% (Figure S2, Table S2). Thus, as long as a candidate vaccine is shown to have a fold change neutralization titer compared to the comparator vaccine that is no less than this margin in a non-inferiority trial (or more than this margin in a superiority trial), then the vaccine efficacy of the candidate vaccine has a high confidence of being above 80%.

**Estimating assay variability for predicting an individual's level of protection**
The 29 replicate estimates of neutralization on the same standard (i.e. WHO international standard 20/130) allow quantification of the precision of an assay's estimate for an individual's neutralization titer. The standard deviation of the $\log_{10}$ neutralization titers

measured in these 29 technical replicates of the 20/130 standard was 0.41 in the above described assay. Assuming an individual has a true neutralization titer of $N$, the chance of observing a neutralization titer $n$ for that individual on repetition of the neutralization assay is described by a normally distributed random variable with mean $N$ and standard deviation 0.41. Therefore, the measurements of the mean of the duplicate assay for such an individual will follow a normal distribution with mean $N$ and standard deviation $0.5 \times \sqrt{2 \times 0.41^2} = 0.29$. It follows that the observed neutralization titer of an individual with a true neutralization titer of $N$ is expected to fall within the range $(N \times 10^{-z_{crit}}, N \times 10^{z_{crit}})$ for 95% of observations, where $z_{crit}$ is the 2.5$^{th}$ percentile of the normal distribution with mean 0 and standard deviation 0.29. These data are used in the main text to determine the precision in the estimated efficacy of an individual given they are observed to have a particular neutralization titer.

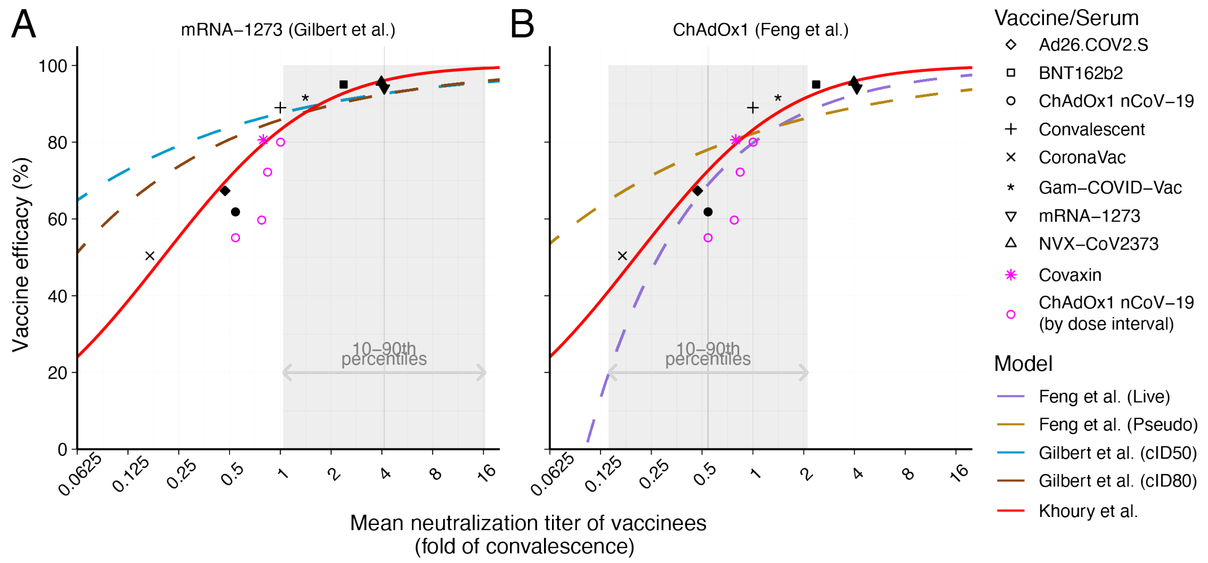


**Figure S1. Predicting the efficacy of other vaccines (immunobridging):** Both the breakthrough-infection (*4, 5*) and vaccine-comparison (*1*) models can be used for 'immunobridging' to estimate the efficacy of novel vaccines (explained in supplementary material), using the protection curve and the distribution of neutralization titers of vaccinees to estimate overall vaccine efficacy. The vaccine-comparison model (red line) is fitted to the data on neutralization and protection for seven individual vaccines and convalescent subjects (black shapes). The predicted vaccine efficacy from the breakthrough infection model applied to mRNA-1273 vaccinees is shown (A). Two different protection curves were derived using either the 50% and 80% in vitro neutralization titers (labeled as ID50 and ID80, respectively)(*5*). The shaded area indicates the 10$^{th}$ - 90$^{th}$ percentiles of the neutralization data. (B) The predicted vaccine efficacy from study of breakthrough infection in ChAdOx1 nCoV19 vaccinees is also shown using neutralization data from either a SARS-CoV-2 (purple) or pseudovirus (light brown) neutralization assays (*4*) (colored rectangle indicates 10-90$^{th}$ percentiles of the data). We see that the breakthrough-infection models agree closely with both the vaccine-comparison model and the reported vaccine efficacies in the ranges where data was available for each study. The pink star indicate reported efficacy of an eighth vaccine(*15*) and pink open circles indicate the results of a meta-analysis of ChAdOx1 nCoV19 at different dose intervals, showing higher efficacy and neutralization titers with wider dose intervals(*11*).

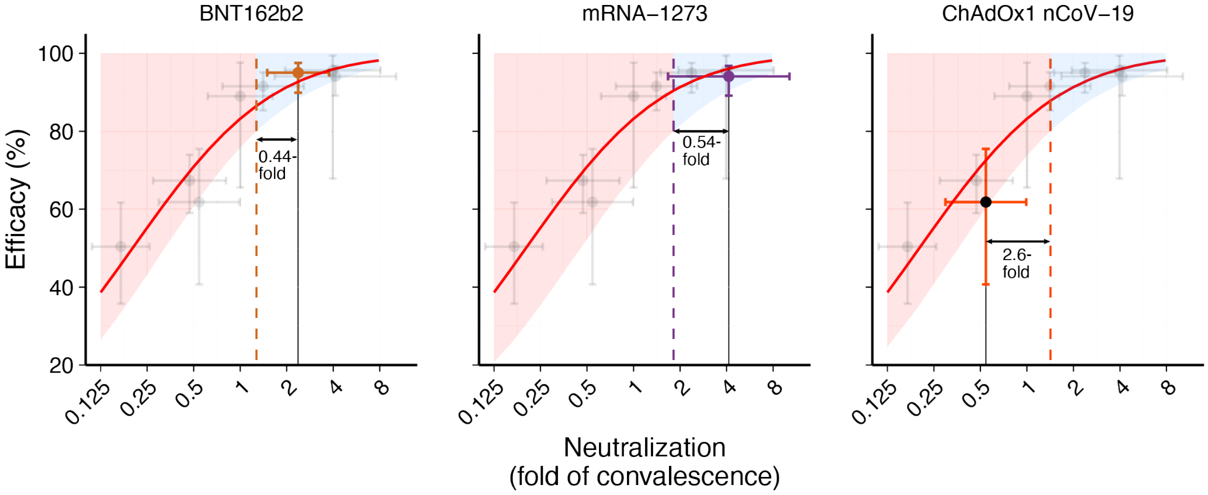


**Figure S2:** The model published by Khoury et al. (*1*) can be used to predict the fold drop in neutralization titer compared to a reference vaccine (either BNT162b2 or mRNA-1273) that would lead to an efficacy estimate where the lower 95% Confidence bound is 80%. The red curve is the model relating neutralizing antibodies and efficacy from Khoury et al. The shaded region indicates the upper 95% confidence region of vaccine efficacy estimates from the Khoury et al. model. The colored dots are the neutralizing antibody titer for each of the three reference vaccines as well as the reported efficacy from the original clinical trials (*7, 9*). The grey dots are the mean neutralizing antibody titers and reported vaccine efficacies of the other vaccines used in fitting the Khoury et al model (*1*). All error bars are 95% confidence intervals. From this model, if a candidate vaccine can be demonstrated to induce a GMT that is more than 0.44-fold (i.e. 44%) of the level seen in BNT162b2 vaccinees, or more than 0.54-fold (i.e. 54%) of the level observed in mRNA-1273 vaccinees, than the Khoury et al. model would predict such a vaccine has an efficacy with a lower 95% confidence interval of 80%. Similarly, the same approach predicts that a candidate vaccine should have a GMT at least 2.6-fold higher the level seen in ChAdOx1 nCoV-19 vaccinees in order that there is high confidence that the predicted efficacy of the candidate vaccine is above 80%. Note that these non-inferiority/superiority margins are computed independently for different reference vaccines and depends on the uncertainty in the model parameters, as well as the uncertainty in the actual position of the reference vaccines (based on the Phase 1/2 clinical trial data) on the fold-convalescence scale (i.e. the horizontal error bars of the reference has been included in the margins reported for each vaccine).

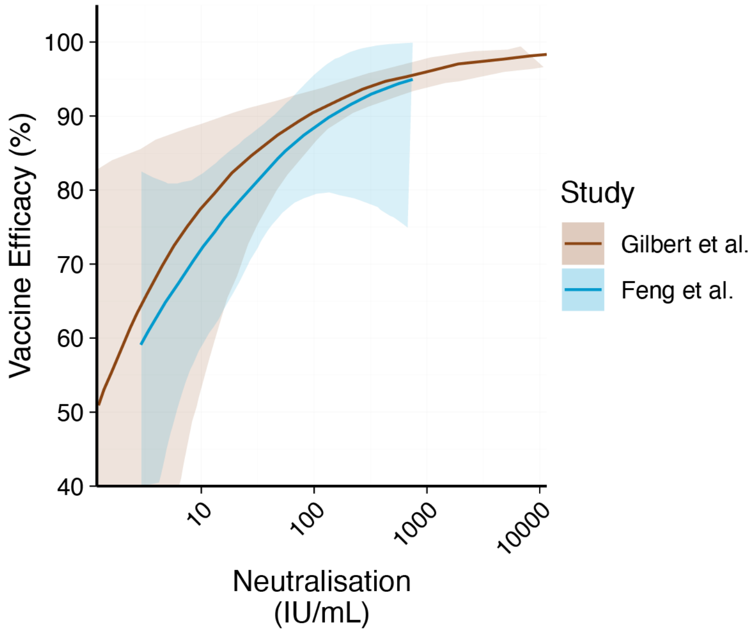


**Figure S3:** The models from Gilbert et al. (brown) (*5*) and Feng et al. (blue) (*4*), showing the estimated relationship between neutralization titer (measured using a pseudo-virus neutralization assay with 50% neutralization endpoint calibrated to international units) and vaccine efficacy. The model and 95% confidence bands were extracted from the respective studies as described in the data extraction section. This highlights that the relationship between neutralization and vaccine efficacy is perhaps more similar when the assay used to define the relationship is more similar between studies.

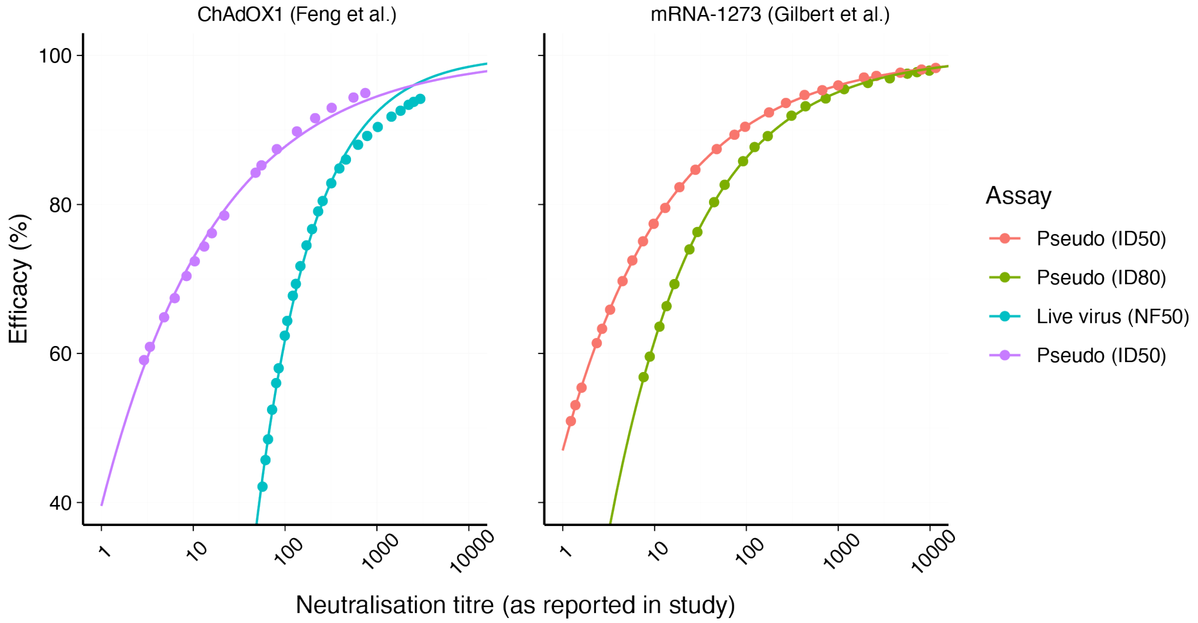


**Figure S4:** Fitting the extracted model (dots) describing the relationship between neutralization titers and protection as reported in (*5*) and (*4*), with the functional form of this relationship (equation 1). Line represents the fitted model in each case. Note that the dots are not evenly spaced as these were extracted manually from the figures using WebPlotDigitizer online application (https://automeris.io/WebPlotDigitizer/). These fits allowed the parameters a, b and c to be estimated for equation 1 (see Table S4).

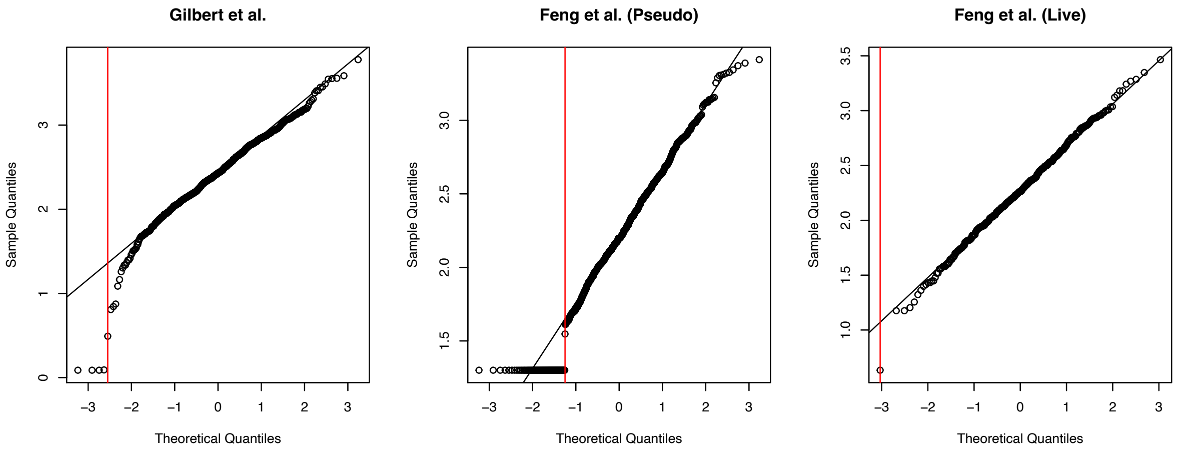


**Figure S5:** Normality of neutralization distributions. Neutralization data from Gilbert ID50 assay was extracted from figure S10 of the supplementary material of(*5*) using the web tool described above and is not as reliable as the data extracted from the Feng et al. study using Adobe Illustrator.

| Study | Cohort size | Timing (gender) | Median Age (range) | Assay | GMT (IU/ml) | Ref |
|---|---|---|---|---|---|---|
| Gilbert et al. | n=1005 | 28 days post second dose (47% Female) | 55 (18-87) | Pseudo-neutralization (50% inhibition) | 247 | (*5*) |
| Huang et al. | N=30 | 28 days post second dose | NR | Pseudo-neutralization (2 methods used, Duke and Monogram) (50% inhibition) | 480 or 275 (Duke and Monogram respectively) | (*16*) |
| Khoury et al. / Jackson et al. | n=15 | 14 days post second dose, (53% Female) | 31 (18-55) | Live-virus neutralization | 1,057 | (*1, 7*) |
| Garcia-Beltran et al. | n=24 | within 3 months of 2nd dose (71% Female) | 54 (24-72) | Pseudo-neutralization (50% inhibition) | 1,362 | (*17*) |
| Zhang et al. | n=30 | Within the first 28 days post second dose (60% Female) | 44 (NR) | Pseudo-neutralization (50% inhibition) | 1,399 | (*18*) |
| Kung et al. | n=20 | 14 days post second dose | NR (22-69) | Live-virus neutralization | 1,404 | (*19*) |

**Table S1: Variation in geometric mean neutralization titers (reported in IU/mL):** Summary of different studies identified that report the mean neutralization titer of a group of mRNA-1273 vaccinees using different assays, and which have all been calibrated to international units (IU/mL). The range of estimates is 247-1404 IU/mL, highlighting that even after calibrating assays to the WHO established international standards discrepancies remain between laboratories and assays.

| Comparator Vaccine | Non-inferiority or Superiority trial | Margin predicted to give at least 80% efficacy (fold-change in the GMT of candidate compared to the comparator vaccine) |
|---|---|---|
| BNT162b2 | Non-inferiority | 0.54 |
| mRNA-1273 | Non-inferiority | 0.44 |
| ChAdOx1 nCoV-19 | Superiority | 2.6 |

**Table S2:** Estimated margins for non-inferiority (or superiority) trials which seek to compare a new vaccine candidate against an existing comparator vaccine. The model reported by Khoury et al. (*1*) predicts that as long as the difference in neutralizing antibody titer between the comparator and reference vaccine is not less than (or is greater than, in the case of a superiority trial) the margin reported here, then the efficacy of the new candidate is predicted to have a lower bound 95% CI of 80%.

| Study | Assay | Estimated standard deviation of ($log_{10}$) neutralization titers in uninfected vaccinees# |
|---|---|---|
| Gilbert et al. | ID50 (Pseudovirus) | 0.47 |
| Gilbert et al. | ID80 (Pseudovirus) | 0.43 |
| Feng et al. | ID50 (Pseudovirus) | 0.46 |
| Feng et al. | NF50 (Live virus) | 0.41 |

**Table S3:** Estimates of SD of neutralization data from each study. #Gilbert et al.(*5*) using the confidence intervals of the reported mean neutralization titer and Feng et al. (*4*) from censored regression.

| Study | Assay | Parameters | | |
|---|---|---|---|---|
| | | a | b | c |
| Gilbert et al. | ID50 (Pseudovirus) | $3.3\times10^{-3}$ | 1.0 | $3.3\times10^{2}$ |
| Gilbert et al. | ID80 (Pseudovirus) | $2.3\times10^{-3}$ | 0.86 | $2.3\times10^{2}$ |
| Feng et al. | ID50 (Pseudovirus) | $2.5\times10^{-3}$ | 0.80 | $2.5\times10^{2}$ |
| Feng et al. | NF50 (Live virus) | $9.8\times10^{-3}$ | 1.2 | $9.8\times10^{2}$ |

**Table S4:** Estimated parameters for equation (1) from fitting risk model in figure S4.

| Study | Assay | Efficacy (%) | Source |
|---|---|---|---|

| | | | |
|---|---|---|---|
| Gilbert et al. | Both ID50 and ID80 (Pseudovirus) | 92.8 | Figure 4 of (*5*) |
| Feng et al. | ID50 (Pseudovirus) | 78.1 | Calculated using equation (2) and parameters in table S3 and S4. |
| | Live virus (NF50) | 69.0 | |
| Bergwerk et al. | Pseudoviral assay | 94.0 | (*12*) |

**Table S5:** Efficacies used for each study to calculate unadjusted protection curves from breakthrough infection data.

**References**


1. D. S. Khoury, D. Cromer, A. Reynaldi, T. E. Schlub, A. K. Wheatley, J. A. Juno, K. Subbarao, S. J. Kent, J. A. Triccas, M. P. Davenport, Neutralizing antibody levels are highly predictive of immune protection from symptomatic SARS-CoV-2 infection. *Nat Med* **27**, 1205-1211 (2021).
2. A. K. Wheatley, J. A. Juno, J. J. Wang, K. J. Selva, A. Reynaldi, H. X. Tan, W. S. Lee, K. M. Wragg, H. G. Kelly, R. Esterbauer, S. K. Davis, H. E. Kent, F. L. Mordant, T. E. Schlub, D. L. Gordon, D. S. Khoury, K. Subbarao, D. Cromer, T. P. Gordon, A. W. Chung, M. P. Davenport, S. J. Kent, Evolution of immune responses to SARS-CoV-2 in mild-moderate COVID-19. *Nat Commun* **12**, 1162 (2021).
3. W. H. Organization, Establishment of the WHO International Standard and Reference Panel for anti-SARS-CoV-2 antibody. *Expert Committee on Biological Standardization, Geneva*, 9-10 (2020).
4. S. Feng, D. J. Phillips, T. White, H. Sayal, P. K. Aley, S. Bibi, C. Dold, M. Fuskova, S. C. Gilbert, I. Hirsch, H. E. Humphries, B. Jepson, E. J. Kelly, E. Plested, K. Shoemaker, K. M. Thomas, J. Vekemans, T. L. Villafana, T. Lambe, A. J. Pollard, M. Voysey, C. V. T. G. Oxford, Correlates of protection against symptomatic and asymptomatic SARS-CoV-2 infection. *Nat Med* **27**, 2032-2040 (2021).
5. P. B. Gilbert, D. C. Montefiori, A. B. McDermott, Y. Fong, D. Benkeser, W. Deng, H. Zhou, C. R. Houchens, K. Martins, L. Jayashankar, F. Castellino, B. Flach, B. C. Lin, S. O'Connell, C. McDanal, A. Eaton, M. Sarzotti-Kelsoe, Y. Lu, C. Yu, B. Borate, L. W. P. v. d. Laan, N. S. Hejazi, C. Huynh, J. Miller, H. M. E. Sahly, L. R. Baden, M. Baron, L. D. L. Cruz, C. Gay, S. Kalams, C. F. Kelley, M. P. Andrasik, J. G. Kublin, L. Corey, K. M. Neuzil, L. N. Carpp, R. Pajon, D. Follmann, R. O. Donis, R. A. Koup, Immune correlates analysis of the mRNA-1273 COVID-19 vaccine efficacy clinical trial. *Science* **375**, 43-50 (2022).
6. M. Bergwerk, T. Gonen, Y. Lustig, S. Amit, M. Lipsitch, C. Cohen, M. Mandelboim, E. G. Levin, C. Rubin, V. Indenbaum, I. Tal, M. Zavitan, N. Zuckerman, A. Bar-Chaim, Y. Kreiss, G. Regev-Yochay, Covid-19 Breakthrough Infections in Vaccinated Health Care Workers. *N Engl J Med* **385**, 1474-1484 (2021).
7. L. A. Jackson, E. J. Anderson, N. G. Rouphael, P. C. Roberts, M. Makhene, R. N. Coler, M. P. McCullough, J. D. Chappell, M. R. Denison, L. J. Stevens, A. J. Pruijssers, A. McDermott, B. Flach, N. A. Doria-Rose, K. S. Corbett, K. M. Morabito, S. O'Dell, S. D. Schmidt, P. A. Swanson, 2nd, M. Padilla, J. R. Mascola, K. M. Neuzil, H. Bennett, W. Sun, E. Peters, M. Makowski, J. Albert, K. Cross, W. Buchanan, R. Pikaart-Tautges, J. E. Ledgerwood, B. S. Graham, J. H. Beigel, R. N. A. S. G. m, An mRNA Vaccine against SARS-CoV-2 - Preliminary Report. *N Engl J Med* **383**, 1920-1931 (2020).
8. E. E. Walsh, R. W. Frenck, Jr., A. R. Falsey, N. Kitchin, J. Absalon, A. Gurtman, S. Lockhart, K. Neuzil, M. J. Mulligan, R. Bailey, K. A. Swanson, P. Li, K. Koury, W. Kalina, D. Cooper, C. Fontes-Garfias, P. Y. Shi, O. Tureci, K. R. Tompkins, K. E. Lyke, V. Raabe, P. R. Dormitzer, K. U. Jansen, U. Sahin, W. C. Gruber, Safety and Immunogenicity of Two RNA-Based Covid-19 Vaccine Candidates. *N Engl J Med* **383**, 2439-2450 (2020).
9. P. M. Folegatti, K. J. Ewer, P. K. Aley, B. Angus, S. Becker, S. Belij-Rammerstorfer, D. Bellamy, S. Bibi, M. Bittaye, E. A. Clutterbuck, C. Dold, S. N. Faust, A. Finn, A. L. Flaxman, B. Hallis, P. Heath, D. Jenkin, R. Lazarus, R. Makinson, A. M. Minassian, K. M. Pollock, M. Ramasamy, H. Robinson, M. Snape, R. Tarrant, M. Voysey, C. Green,

A. D. Douglas, A. V. S. Hill, T. Lambe, S. C. Gilbert, A. J. Pollard, C. V. T. G. Oxford, Safety and immunogenicity of the ChAdOx1 nCoV-19 vaccine against SARS-CoV-2: a preliminary report of a phase 1/2, single-blind, randomised controlled trial. *Lancet* **396**, 467-478 (2020).

10. M. Voysey, S. A. C. Clemens, S. A. Madhi, L. Y. Weckx, P. M. Folegatti, P. K. Aley, B. Angus, V. L. Baillie, S. L. Barnabas, Q. E. Bhorat, S. Bibi, C. Briner, P. Cicconi, A. M. Collins, R. Colin-Jones, C. L. Cutland, T. C. Darton, K. Dheda, C. J. A. Duncan, K. R. W. Emary, K. J. Ewer, L. Fairlie, S. N. Faust, S. Feng, D. M. Ferreira, A. Finn, A. L. Goodman, C. M. Green, C. A. Green, P. T. Heath, C. Hill, H. Hill, I. Hirsch, S. H. C. Hodgson, A. Izu, S. Jackson, D. Jenkin, C. C. D. Joe, S. Kerridge, A. Koen, G. Kwatra, R. Lazarus, A. M. Lawrie, A. Lelliott, V. Libri, P. J. Lillie, R. Mallory, A. V. A. Mendes, E. P. Milan, A. M. Minassian, A. McGregor, H. Morrison, Y. F. Mujadidi, A. Nana, P. J. O'Reilly, S. D. Padayachee, A. Pittella, E. Plested, K. M. Pollock, M. N. Ramasamy, S. Rhead, A. V. Schwarzbold, N. Singh, A. Smith, R. Song, M. D. Snape, E. Sprinz, R. K. Sutherland, R. Tarrant, E. C. Thomson, M. E. Torok, M. Toshner, D. P. J. Turner, J. Vekemans, T. L. Villafana, M. E. E. Watson, C. J. Williams, A. D. Douglas, A. V. S. Hill, T. Lambe, S. C. Gilbert, A. J. Pollard, C. V. T. G. Oxford, Safety and efficacy of the ChAdOx1 nCoV-19 vaccine (AZD1222) against SARS-CoV-2: an interim analysis of four randomised controlled trials in Brazil, South Africa, and the UK. *Lancet* **397**, 99-111 (2021).
11. M. Voysey, S. A. Costa Clemens, S. A. Madhi, L. Y. Weckx, P. M. Folegatti, P. K. Aley, B. Angus, V. L. Baillie, S. L. Barnabas, Q. E. Bhorat, S. Bibi, C. Briner, P. Cicconi, E. A. Clutterbuck, A. M. Collins, C. L. Cutland, T. C. Darton, K. Dheda, C. Dold, C. J. A. Duncan, K. R. W. Emary, K. J. Ewer, A. Flaxman, L. Fairlie, S. N. Faust, S. Feng, D. M. Ferreira, A. Finn, E. Galiza, A. L. Goodman, C. M. Green, C. A. Green, M. Greenland, C. Hill, H. C. Hill, I. Hirsch, A. Izu, D. Jenkin, C. C. D. Joe, S. Kerridge, A. Koen, G. Kwatra, R. Lazarus, V. Libri, P. J. Lillie, N. G. Marchevsky, R. P. Marshall, A. V. A. Mendes, E. P. Milan, A. M. Minassian, A. McGregor, Y. F. Mujadidi, A. Nana, S. D. Padayachee, D. J. Phillips, A. Pittella, E. Plested, K. M. Pollock, M. N. Ramasamy, A. J. Ritchie, H. Robinson, A. V. Schwarzbold, A. Smith, R. Song, M. D. Snape, E. Sprinz, R. K. Sutherland, E. C. Thomson, M. E. Torok, M. Toshner, D. P. J. Turner, J. Vekemans, T. L. Villafana, T. White, C. J. Williams, A. D. Douglas, A. V. S. Hill, T. Lambe, S. C. Gilbert, A. J. Pollard, C. V. T. G. Oxford, Single-dose administration and the influence of the timing of the booster dose on immunogenicity and efficacy of ChAdOx1 nCoV-19 (AZD1222) vaccine: a pooled analysis of four randomised trials. *Lancet* **397**, 881-891 (2021).
12. G. Chodick, L. Tene, R. S. Rotem, T. Patalon, S. Gazit, A. Ben-Tov, C. Weil, I. Goldshtein, G. Twig, D. Cohen, K. Muhsen, The effectiveness of the two-dose BNT162b2 vaccine: analysis of real-world data. *Clin Infect Dis* **74**, 472-478 (2022).
13. P. B. Gilbert, Y. Fong, M. Carone, Assessment of immune correlates of protection via controlled vaccine efficacy and controlled risk. *arXiv preprint arXiv:2107.05734*, (2021).
14. D. Cromer, M. Steain, A. Reynaldi, T. E. Schlub, A. K. Wheatley, J. A. Juno, S. J. Kent, J. A. Triccas, D. S. Khoury, M. P. Davenport, Neutralising antibody titres as predictors of protection against SARS-CoV-2 variants and the impact of boosting: a meta-analysis. *The Lancet Microbe* **3**, e52-61 (2021).

15. R. Ella, S. Reddy, W. Blackwelder, V. Potdar, P. Yadav, V. Sarangi, V. K. Aileni, S. Kanungo, S. Rai, P. Reddy, S. Verma, C. Singh, S. Redkar, S. Mohapatra, A. Pandey, P. Ranganadin, R. Gumashta, M. Multani, S. Mohammad, P. Bhatt, L. Kumari, G. Sapkal, N. Gupta, P. Abraham, S. Panda, S. Prasad, B. Bhargava, K. Ella, K. M. Vadrevu, C. S. Group, Efficacy, safety, and lot-to-lot immunogenicity of an inactivated SARS-CoV-2 vaccine (BBV152): interim results of a randomised, double-blind, controlled, phase 3 trial. *Lancet* **398**, 2173-2184 (2021).
16. Y. Huang, O. Borisov, J. J. Kee, L. N. Carpp, T. Wrin, S. Cai, M. Sarzotti-Kelsoe, C. McDanal, A. Eaton, R. Pajon, J. Hural, C. M. Posavad, K. Gill, S. Karuna, L. Corey, M. J. McElrath, P. B. Gilbert, C. J. Petropoulos, D. C. Montefiori, Calibration of two validated SARS-CoV-2 pseudovirus neutralization assays for COVID-19 vaccine evaluation. *Sci Rep* **11**, 23921 (2021).
17. W. F. Garcia-Beltran, K. J. St Denis, A. Hoelzemer, E. C. Lam, A. D. Nitido, M. L. Sheehan, C. Berrios, O. Ofoman, C. C. Chang, B. M. Hauser, J. Feldman, A. L. Roederer, D. J. Gregory, M. C. Poznansky, A. G. Schmidt, A. J. Iafrate, V. Naranbhai, A. B. Balazs, mRNA-based COVID-19 vaccine boosters induce neutralizing immunity against SARS-CoV-2 Omicron variant. *Cell* **185**, 457-466 e454 (2022).
18. Z. Zhang, J. Mateus, C. H. Coelho, J. M. Dan, C. R. Moderbacher, R. I. Gálvez, F. H. Cortes, A. Grifoni, A. Tarke, J. Chang, E. A. Escarrega, C. Kim, B. Goodwin, N. I. Bloom, A. Frazier, D. Weiskopf, A. Sette, S. Crotty, Humoral and cellular immune memory to four COVID-19 vaccines. *bioRxiv*, 2022.2003.2018.484953 (2022).
19. Y.-A. Kung, C.-G. Huang, S.-Y. Huang, K.-T. Liu, P.-N. Huang, K.-Y. Yu, S.-L. Yang, C.-P. Chen, C.-Y. Cheng, Y.-T. Lin, Y.-C. Liu, G.-W. Chen, S.-R. Shih, Antibody titers measured by commercial assays are correlated with neutralizing antibody titers calibrated by international standards. *medRxiv*, 2021.2007.2016.21260618 (2021).
